# Socioeconomic, demographic and environmental factors inform intervention prioritization in urban Nigeria

**DOI:** 10.1101/2022.03.16.22272476

**Authors:** Chilochibi Chiziba, Ousmane Diallo, Amelia Bertozzi-Villa, Dan Weiss, Laina Mercer, Jaline Gerardin, Ifeoma D. Ozodiegwu

## Abstract

Nigeria is one of three countries projected to have the largest absolute increase in the size of its urban population and this could intensify malaria transmission in cities. Accelerated urban population growth is out-pacing the availability of affordable housing and basic services and resulting in living conditions that foster vector breeding and heterogeneous malaria transmission. Understanding community determinants of malaria transmission in urban areas informs the targeting of interventions to populations at greatest risk. In this study, we analyzed cluster-level data from the Demographic and Health Surveys (DHS) and the Malaria Indicator Survey (MIS) as well as geospatial covariates to describe malaria burden and its determinants in areas administratively defined as urban in Nigeria. Overall, we found low malaria test positivity across urban areas,. We observed declines in test positivity rate over time and identified the percentage of individuals with post-primary education, the percentage of individuals in the rich wealth quintiles, the percentage of individuals living in improved housing in 2015, all age population density, median age, the percentage of children under the age of five that sought medical treatment for fever, total precipitation and enhanced vegetation index as key community predictors of malaria transmission intensity. The unrepresentativeness of the DHS and MIS in urban settings at the state and geopolitical zonal level, regional differences in malaria seasonality across Nigeria, and information detection bias were among likely factors that limited our ability to compare malaria burden across geographic space and ultimately drove model uncertainty. Nevertheless, study findings provide a starting point for informing decisions on intervention prioritization within urban spaces and underscore the need for improved regionally-focused surveillance systems in Nigeria.

## Introduction

Nigeria accounts for 27 percent of all global malaria cases and deaths, respectively, making it the greatest contributor to the global malaria burden [1] Underlying Nigeria’s malaria burden are spatial and temporal differences in malaria risk driven by variations in ecological and climatic factors, intervention histories, health system factors, land-use, and urbanization [2–4]. Nigeria is one of three countries expected to account for nearly one-third of global urban population growth between 2018 and 2050 [5]. Half of Nigeria’s roughly 200 million population lived in urban areas in 2018 and the proportion of urban residents is projected to increase to 70 percent by 2050 [5]. Although urbanization is expected to decrease malaria transmission by reducing natural vector breeding sites and biting risk [6–8], rapid and unplanned urbanization characterized by the siting of farms within urban neighborhoods and the development and expansion of informal settlements with inadequate basic services and poor sanitary conditions support the creation of natural and artficial habitats for larval development [9–11]. In addition, urban settlers, particularly those of lower socio-economic status, maintain rural residences and connections and the resultant urban-rural mobility could introduce parasites from rural villages, sustaining transmission in urban spaces [12–14]. The impact of environmental and anthropogenic factors are often reflected in city-level heterogeneities in malaria risk [15], prompting the need to prioritize interventions in areas of greatest need as well as deprioritize interventions in areas of minimal risk. Intervention targeting would ideally be achieved by stratifying localities using epidemiological, ecological, health system and socio-economic determinants following the World Health Organization’s recommendations [15], but information on how these determinants relate to malaria transmission, to facilitate locality grouping in urban Nigeria is lacking.

Community determinants of malaria transmission in urban Nigeria are not well-researched. Most studies examine only individual level factors [16–19], are limited to specific urban settlement archetypes [17,19] or are hospital-based [17,20]. In a study of patients receiving care at two hospitals in the city of Ibadan, Awosolu and colleagues found that distance from streams within 1km and travel to a rural area in the last months were statistically significant risk factors for malaria infection, however, participants’ place of residence was not accounted for, increasing the likelihood of inaccurate results [20]. Findings from a cross-sectional survey in an urban town in Nigeria’s South-West region indicated that the types of windows and environmental hygiene significantly predicted malaria prevalence within households but this study provided no information on categorization of these covariates and their measures of association [19]. Insufficient information and the methodological gaps in the extant literature motivate the need for additional studies that elucidate major drivers of malaria risk in urban Nigeria.

Georeferenced survey data, such as the Malaria Indicator Surveys (MIS) and Demographic and Health surveys (DHS), and modeled geospatial data can be utilized to understand urban malaria transmission risk. Unlike routine surveillance systems that collect solely information on malaria infection status and are biased towards individuals that live close to health facilities or seek care in public institutions [21,22], the MIS and DHS uses a cluster sampling methodology to collect data on individual level infection status and risk factors that can be aggregated for urban clusters, and when supplemented by geospatial rasters facilitate examination of risk factor associations. Moreover, definitions of urban areas within the DHS and MIS survey are aligned with those of local administrators increasing the likelihood that analysis results will be accepted by policymakers. Although DHS and MIS data is too sparse to characterize malaria risk within cities, understanding transmission risk between different urban settings can provide a level of insight.

Therefore, in light of data limitations and the need to understand how to appropriately group geographic areas in Nigeria for intervention targeting, this study analyzed DHS, MIS and geospatial data to 1) quantify spatio-temporal variation in malaria test positivity rate among children under the age of five years (U5), 2) identify predictors of U5 malaria test positivity rate, and 3) generate model effect plots to describe associations between covariates and U5 malaria test positivity rate. Predicted effects observed in unadjusted effect plots represent correlations between dependent and independent variables that could be leveraged to choose potential thresholds for intervention prioritization. Adjusted effects provide insight into the potential causal impact of independent variables, and are essential for understanding the public health impact of interventions.

## Methods

### Data

Cluster-level data from the 2010 and 2015 MIS, 2018 DHS and publicly available geospatial malaria covariate data were used for this study (see Table 1 for references). The MIS and DHS are complex multistage surveys that collect and disseminate accurate, nationally representative data on health and population in over 90 countries [23]. MIS and DHS clusters or enumeration areas are an aggregation of households combined together to facilitate sampling [23]. Surveys are conducted at the individual level after probability proportional to size sampling of clusters and random sampling of households within clusters [23]. For purposes of this study, only clusters classified as located in urban areas were retained [24]. GPS coordinates of urban clusters are displaced by a distance of 0 – 2 km to protect the confidentiality of the survey participants [25]. The number of positive malaria tests by microscopy and the testing sample population for children 6 – 59 months, were summarized for each cluster. Covariate selection was informed by the research literature, which showed that socio-economic, demographic, behavioral, accessibility and environment-related factors are potential explanatory factors for malaria [2,4,26–30]. A description of covariates considered is provided in Table 1. To account for the distance displacement of MIS and DHS clusters during computation of values for geospatial covariates, raster values were aggregated across buffers of up to 4km placed on each MIS and DHS cluster.

**Table 1:** Variable names, definitions and source for cluster-level variables computed from Demographic and Health Surveys

| Variable name | Variable definition | Source |
| --- | --- | --- |
| **Dependent variable** |  |  |
| Number of U5 positive malaria tests | The number of positive malaria tests by microscopy among children 6 – 59 months old aggregated per cluster | MIS and DHS [24,34,35] |
| **Explanatory variables by thematic group** |  |  |
| **Socio-economic factors** |  |  |
| 1. % with post-primary education | Percentage (%) of cluster population with secondary or higher educational attainment | MIS and DHS [24,34,35] |
| 2. % in the rich wealth quintiles | % of cluster population in the rich and richest wealth quintiles. Wealth quintiles were constructed using various indicators of household living standard [36] | MIS and DHS [24,34,35] |
| 3. % in homes with improved flooring | % of cluster population living in homes with improved flooring (finished floors, parquet or polished wood, ceramic tiles, cement and carpet) | MIS and DHS [24,34,35] |
| 4. % in homes with metal or zinc roof | % of cluster population living in homes with a metal or zinc roof | MIS and DHS [24,34,35] |
| 5. % in homes with improved wall type | % of cluster population living in homes with a improved wall type (finished wall, cement, bricks, cement blocks, covered adobe) | MIS and DHS [24,34,35] |
| 6. % living in improved housing (2000) | Predicted % of the cluster population living in improved housing in 2000. Improved housing is defined as homes with improved water and sanitation, sufficient living area and durable construction according to Tusting et al [37]. | Malaria Atlas Project (MAP) [37,38] |
| 7. % living in improved housing (2015) | Predicted % of the cluster population living in improved housing in 2015 | MAP [37,38] |
| *Demographic factors* |  |  |
| 8. All age population density | Estimated population density per cluster at the time of the 2010 and 2015 DHS/MIS surveys. Population density data for 2020 was extracted for the 2018 survey (UN World Population Prospects-Adjusted Population Density, v4.11). Unit is persons per square kilometer | Center for International Earth Science Information Network, Columbia University [39] |
| 9. Population density, children five years and under | Estimated population density for children under the age of five in 2020. Unit is number of children per square kilometer | Humanitarian Data Exchange [40] |
| 10. % of pregnant women | % of pregnant women | MIS and DHS [24,34,35] |
| 11. % of female population | % of females per cluster | MIS and DHS [24,34,35] |
| 12. Median household size | Median household size per cluster | MIS and DHS [24,34,35] |
| 13. Median age | Median age per cluster | MIS and DHS [24,34,35] |
| 14. State | State where the cluster is located | MIS and DHS [24,34,35] |
| 15. Region | Geopolitical region where the cluster is located (Geopolitical regions in Nigeria are six – North East, North West, North Central, South East, South West, South South) | MIS and DHS [24,34,35] |
| *Behavioral factors* |  |  |
| 16. % of individuals using bednets | % of cluster population that slept under a treated bednet the night before the survey | MIS and DHS [24,34,35] |
| 17. % of children 6 – 59 months using bednets, among those tested by microscopy | % of children 6 – 59 months tested for malaria by microscopy that slept under a treated bednet the night before the survey | MIS and DHS [24,34,35] |
| 18. % of U5 children that sought medical treatment for fever | % of children under the age of five that received medical treatment given that they had fever or cough in the two weeks before the survey. Medical treatment must be received in the public sector or medical private sector except for pharmacy | MIS and DHS [24,34,35] |
| 19. % of U5 children with fever that received an artemisinin-combination therapy (ACT) | % of children under the age of five that received an ACT given that they had fever | MIS and DHS [24,34,35] |
| *Accessibility-related factors* |  |  |
| 20. Motorized travel time to health care in minutes | Predicted travel time to healthcare facility in minutes in 2019 | MAP [41] |
| *Environmental factors* |  |  |
| 21. Total precipitation (meters Depth) | Estimated total precipitation during survey month and year per cluster. Units is depth in meters. It is measured as the depth water would have if it is spread evenly over a grid box. | European Center for Medium Range Weather Forecasts (ECMWF), Climate Data Store [42] |
| 22. Temperature (°C) | Estimated temperature of air at 2m above the surface of land, sea or in-land waters in Celsius per cluster during the survey month | ECMWF, Climate Data Store [42] |
| 23. Surface soil moisture (GSM) | Estimated depth averaged amount of water present in a specific soil layer beneath the surface measured as gravimetric soil moisture (GSM) per cluster. GSM is the | Goddard Earth Sciences Data and Information Services Center [43] |
|  | mass of water compared to the mass of solid materials per unit volume of soil |  |
| 24. Distance to water bodies (meters) | Straight line distance to water bodies in meters | MAP (unpublished data) |
| 25. Elevation (meters) | Cluster elevation above sea level in meters | Multi-Error-Removed Improved-Terrain DEM [44] |
| 26. Enhanced Vegetation Index | Enhanced vegetation index for quantifying vegetation greenness in units of spectral index | MAP gap-filled EVI [44] |
| **Other adjustment variables** |  |  |
| 1. Number of children tested | Number of children 6 – 59 months old tested for malaria per cluster | MIS and DHS [24,34,35] |
| 2. Interview date | Year the DHS survey was conducted per cluster and survey month per cluster (some clusters were surveyed over a two month period, the first interview month and associated metric was used) | MIS and DHS [24,34,35] |
| 3. Longitude and latitude | Longitude and latitude positions where the clusters were geolocated after displacement to protect participant confidentiality | MIS and DHS [24,34,35] |

### Descriptive analysis and covariate selection

To understand the burden of U5 malaria across geographic space and time, descriptive analysis of U5 test positivity rate, that is the number U5 positive malaria test divided by the number of U5 children tested for malaria in each cluster, were mapped by state and compared by month and year of survey. This was followed by summaries of covariates to evaluate their distribution and detect likely information bias. Pearson coorelation coefficients were computed by thematic group (Table 1) to identify correlated covariates. The ggplot2 package in R [31] was used for fitting a poisson regression model used to visually assess bivariate relationships between covariates and malaria test positivity. Non-linear relationships with covariates were estimated using natural cubic splines from the splines package in R [32]. When two covariates had a 60% or greater correlation coefficient, the one with the weakest visual relationship with malaria test positivity rate, and wider confidence bands, was not considered in the multivariable analysis.

### Multivariable modeling

A model of U5 malaria test positivity rate was constructed to identify predictors and to generate effect plots. Multivariable generalized linear models were constructed by thematic group and by a combination of variables across thematic group with the glmmTMB package accounting for zero-inflation, temporal and spatial dependence. The dependent variable was the number of positive malaria tests among U5 adjusted for the number of U5 children tested for malaria per cluster using an offset term. Similar to the descriptive models described above, non-linear relationships were modeled using natural cubic splines. Temporal dependence was accounted by modeling the survey month and year for each cluster while the effect of spatial dependence was considered in the model by including the geographic coordinates of each cluster. The Akaike Information Criterion (AIC) statistic was used to select the best predictive model of malaria test positivity rate. The final model was a zero-inflated poisson model of the form below, chosen on the basis of goodness of fit tests (Kolmogorov–Smirnov test, dispersion test, and outlier tests) from the DHARMa package, which uses simulation-based approaches to create interpretable scaled residuals for fitted generalized linear mixed models [33]:

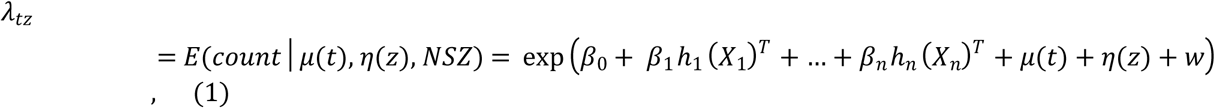

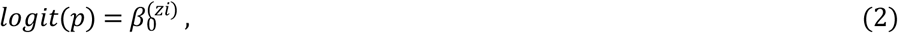

where each of the *β*_*j*_ are vectors of coefficients multiplying their associated vector natural spline basis functions *h*_*j*_ and *β*_0_ represents the model intercept; *X*_*i*_*‥n* are covariate values; *μ*(*t*) is a cluster specific stationary AR(1) process (type of autoregressive model) for modeling temporal dependence by month and year of survey, *t*, for each study cluster ; *η*(*z*) is spatial Matern process for modeling spatial random effects using each cluster coordinate, *z*; NSZ is the event non-structural zero; *w* represents the offset term which is the number of children, 6 – 59, years tested for malaria and the zero components were modeled with the equation in (2) with *p* representing the probability of observing zero counts. Due to the large computational power required to generate cubic plots from complex models, unadjusted and adjusted effect plots for the final model covariates were produced and described using linear splines. All code written in support of this manuscript is available via this doi: 10.5281/zenodo.6350331

### Ethics statement

This study used publicly available, de-identfied data from the 2010 and 2015 MIS and 2018 DHS Surveys; https://dhsprogram.com/Countries/Country-Main.cfm?ctry_id=30&c=Nigeria. Ethical clearance for the MIS and DHS surveys protocols were obtained from the National Health Research Ethics Committee of Nigeria (NHREC) and ICF International’s Institutional Review Boards (IRBs) [24]. The survey ensured compliance with international ethical standards of informed consent and voluntary participation, and privacy and confidentiality. The dataset was requested and a letter of data authorization was received from ICF International through the DHS Program. More details regarding ethical standards of the DHS data are available at: https://www.dhsprogram.com/What-We-Do/Protecting-the-Privacy-of-DHS-Survey-Respondents.cfm.

## Results

### Describing spatial-temporal variation in malaria test positivity in urban areas

#### Sample overview

A total of 794 clusters were sampled in 2010, 2015 and 2018 DHS and MIS surveys. The total number of all age individuals surveyed within each cluster had a wide range in all surveys, from 98 to 2,949 in the 2010 survey, 167 to 2,954 in the 2015 survey, and 3 to 3,471 in the 2018 survey. Malaria test results by microscopy were reported for children 6 – 59 months in 777 of the 794 sampled clusters. The study data consisted of 81 clusters from the 2010 survey, 136 clusters from 2015, and 560 clusters from 2018 (Figure 1a). On average, more children were tested per cluster in 2010 and 2015 compared to 2018 (Figure 1b). The 2010 clusters were sampled in the months of October, November and December. The 2015 clusters were sampled in October and November alone whereas sampling for the 2018 clusters began in August and ended in December.

**Figure 1.**
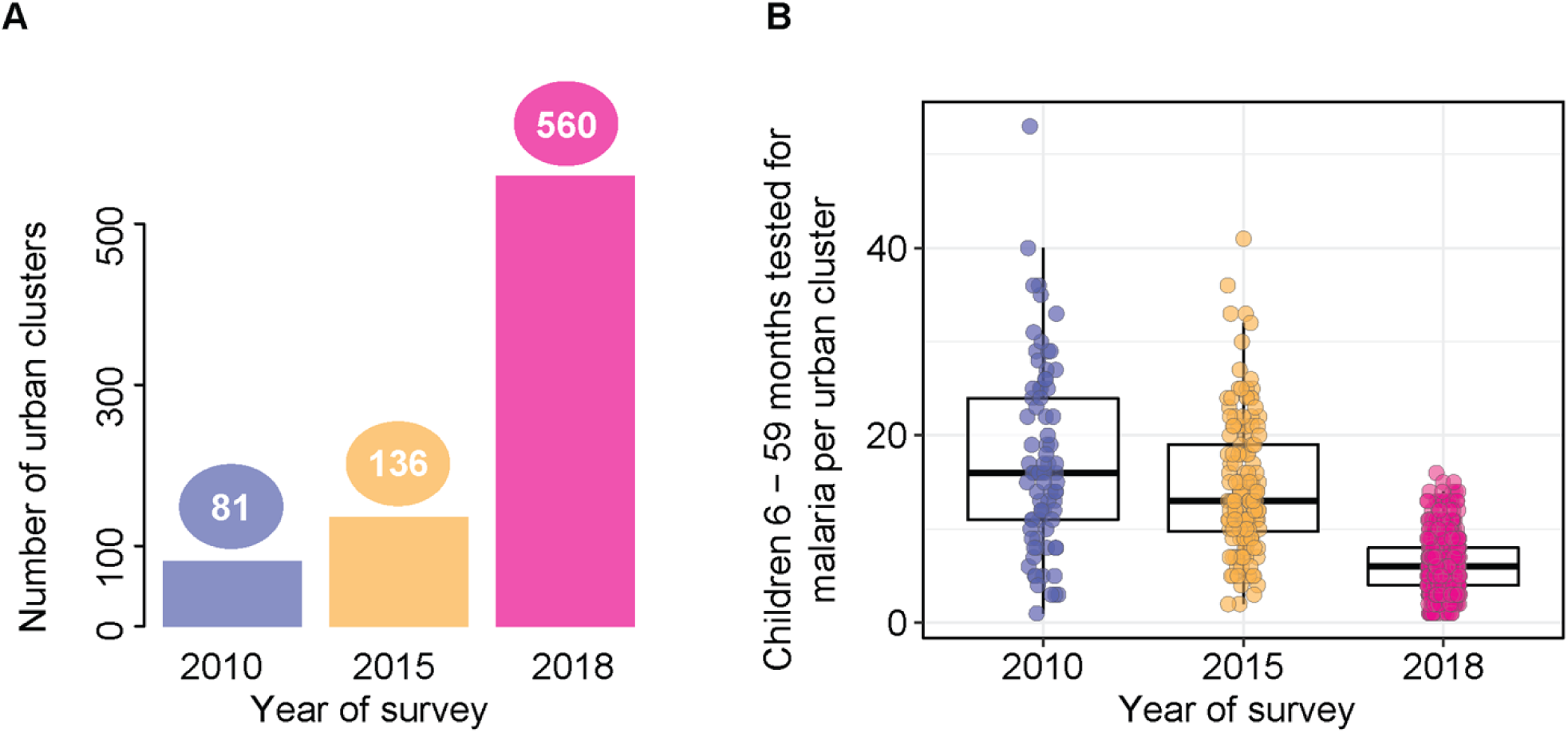
Urban clusters sampled in the 2010, 2015 and 2018 survey. A) Number of clusters per survey year. Eighty-one clusters were sampled in 2010, 136 in 2015 and 560 in 2018. B) Number of children 6 – 59 months tested for malaria per cluster and by year of survey.

#### Low malaria test positivity across majority of urban clusters

An average of nine children (mean = 8.7, standard deviation (SD) = 6.6) were tested per cluster (Figure 2a). The mean number of positive malaria tests per cluster was 1.3 (SD = 2.3). No child tested positive for malaria in 54% (417) of clusters. The median test positivity rate was zero (interquartile range (IQR): 0.2). Stratifying clusters by year of survey revealed that the median test positivity rate was 0.1 for those surveyed in 2010 and 2015, and zero in 2018, and that test positivity rate declined over time (Figure 2b). The majority of clusters in Lagos (90%), Borno (84%) and Akwa Ibom (83%) had a zero test positivity rate (Figure 2c). However, the DHS 2018 report states that 11 LGAs in Borno were dropped during initial sampling due to insecurity [45], implying that the distribution of test positivity rate may not be representative. At the regional level, 66% of clusters in the North East geopolitical region had a zero test positivity rate, 63% in the South-South, 62% in the North-Central, 50% in the South-East, 49% in the South-West and 41% in the North-West (Figure 2d).

**Figure 2:**
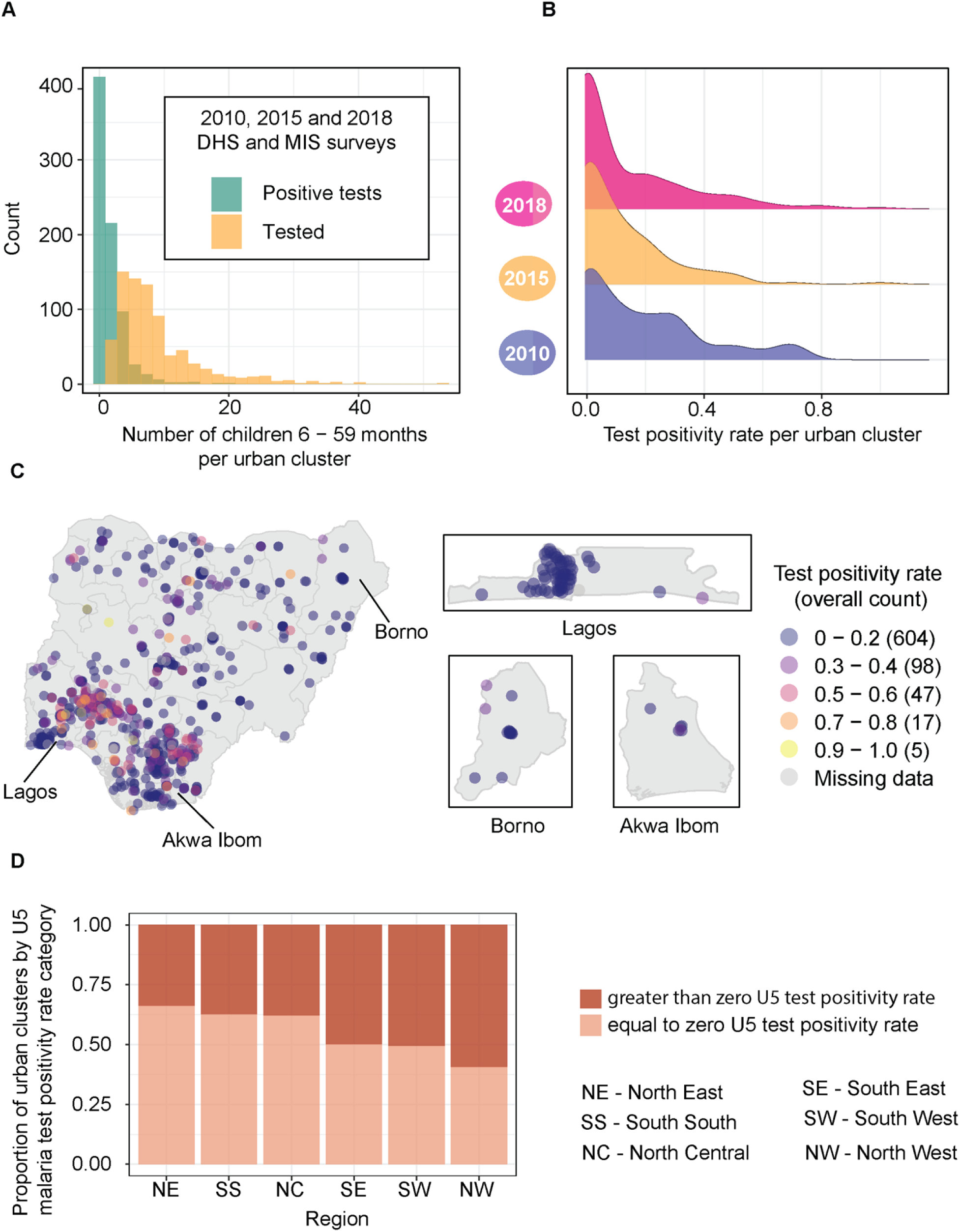
Distribution of malaria tests by microscopy, positive tests and test positivity rates among children 6 – 59 months within urban clusters in the DHS 2010 - 2018. A) Number of malaria tests conducted by microscopy per cluster (yellow), mean = 8.7 (SD = 6.6), and the number tested positive per cluster (green), mean = 1.3 (SD = 2.3). Fifty-four percent (417) of the 777 clusters with non-missing values had zero positive tests. B) Density distribution of cluster test positivity rate by year of DHS survey. C) Positive tests as a fraction of the number of children tested geo-located within state-level geographical boundaries. Majority of surveyed clusters in Lagos, Borno and Akwa Ibom states had zero test positivity rate. D) Regional differences in the proportion of clusters at and above zero malaria test positivity rate

#### Test positivity rates in sampled clusters declined over time

To understand whether the observed temporal declines in test positivity rate in Figure 2b were due to differences in the months during which the 2010, 2015 and 2018 surveys were conducted, clusters sampled in the same months but surveyed in different years were compared. Analysis of 545 clusters for the months of October and November with all three years of survey data, and for December with two years of data, showed that test positivity rates across clusters declined over time, supporting findings observed in Figure 2b. Fifty-three percent of clusters sampled in October had zero positive tests in the 2018 survey as compared to 50% of clusters from the 2015 survey and 32% of clusters from the 2010 survey (Figure 3a). For clusters sampled in November, 59% of clusters from 2018 had zero positive tests compared to 46% and 43% of clusters in 2015 and 2010, respectively (Figure 3b). Similarly, 66% of clusters sampled in December in 2018 in contrast to 13% of clusters in 2010 had zero positive tests (Figure 3c).

**Figure 3.**
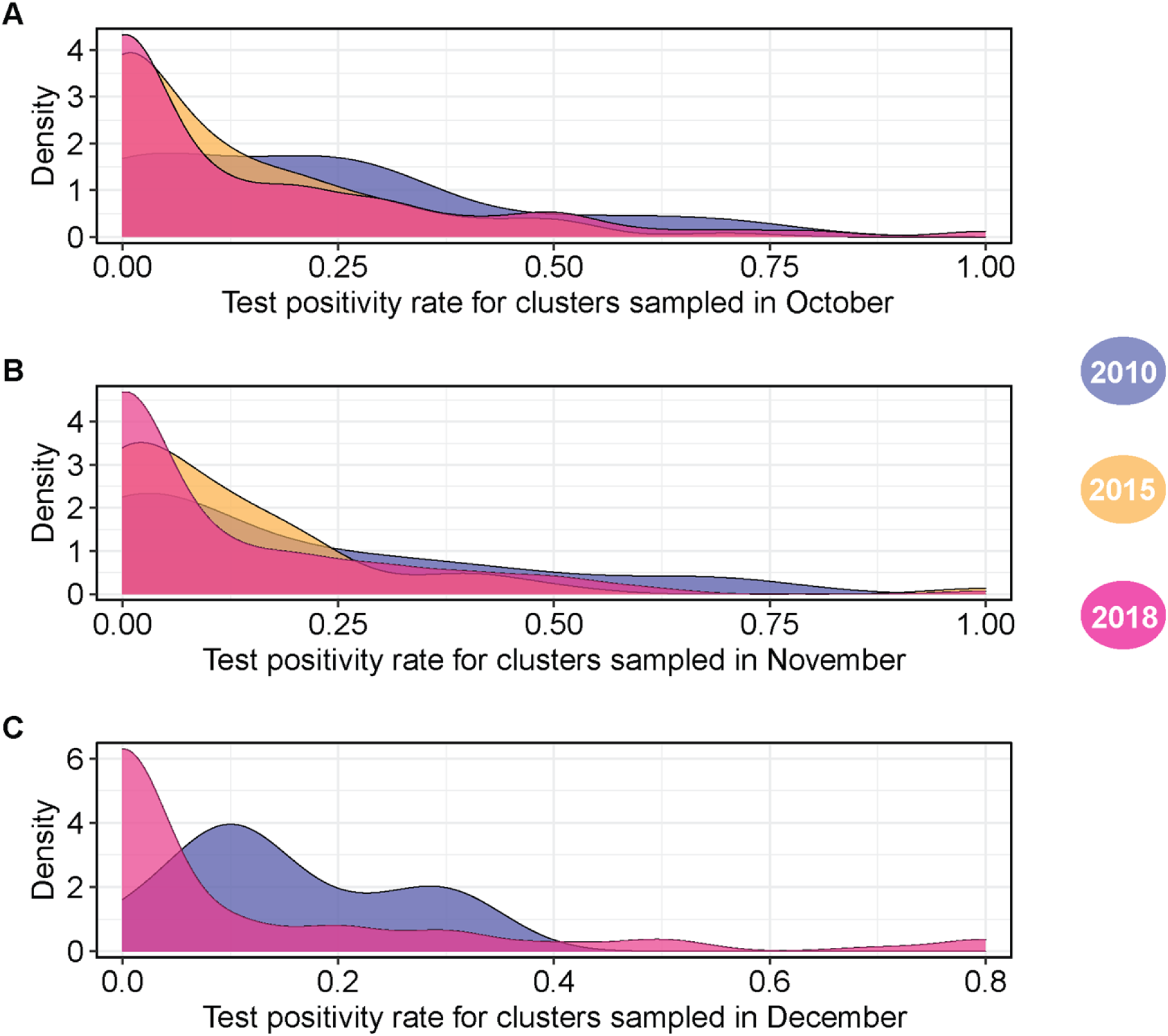
Distribution of test positivity rates by DHS survey year and month. Plots are colored by survey year. The number of children 6 – 59 tested across all clusters and survey years was 253 in October (A), 205 in November (B) and 87 in December (C).

### Identifying predictors of malaria test positivity and visualizing bivariate associations to inform intervention prioritization

Bivariate analysis enabled understanding of the unadjusted functional forms of the relationship between the number of malaria positives and all 26 covariates and informed variable selection for the multivariable regression for highly correlated variables, defined as those with a 60% or greater correlation coefficient. See supplement for visualizations of the distribution of individual covariates, the results of the correlation and bivariate analysis.

The final multivariable prediction model of malaria test positivity was a poisson model with the lowest AIC values was chosen from among a series of 21 models constructed using a combination of model covariates. The model QQ plot showed that model predictions closely approximated the poisson distribution (Supplementary figure 20a). The final model covariates were percentage of individuals with post-primary education, percentage of individuals in the rich wealth quintiles, percentage of individuals living in improved housing in 2015, all age population density, median age, percentage of children under the age of five that sought medical treatment for fever, total precipitation and enhanced vegetation index.

#### Clusters with the lowest educational attainment and wealth were at highest risk for malaria

Socioeconomic variables were negatively associated with malaria transmission intensity, although the effect was less pronounced and more uncertain in the multivariate analysis (Figure 4). Malaria test posivity rate declined with increases in the percentage of individuals with post-primary education. However, the effect of educational attainment flattens out beyond 20% in the unadjusted analysis (Figure 4a). Declines in malaria test positivity rate were also observed with increases in the percentage of individuals in the rich wealth quintiles, particularly between 0 and 50%, and 80 – 100%. In both the adjusted and unadjusted analysis, clusters in the lowest socioeconomic status were at highest risk for malaria.

**Figure 4:**
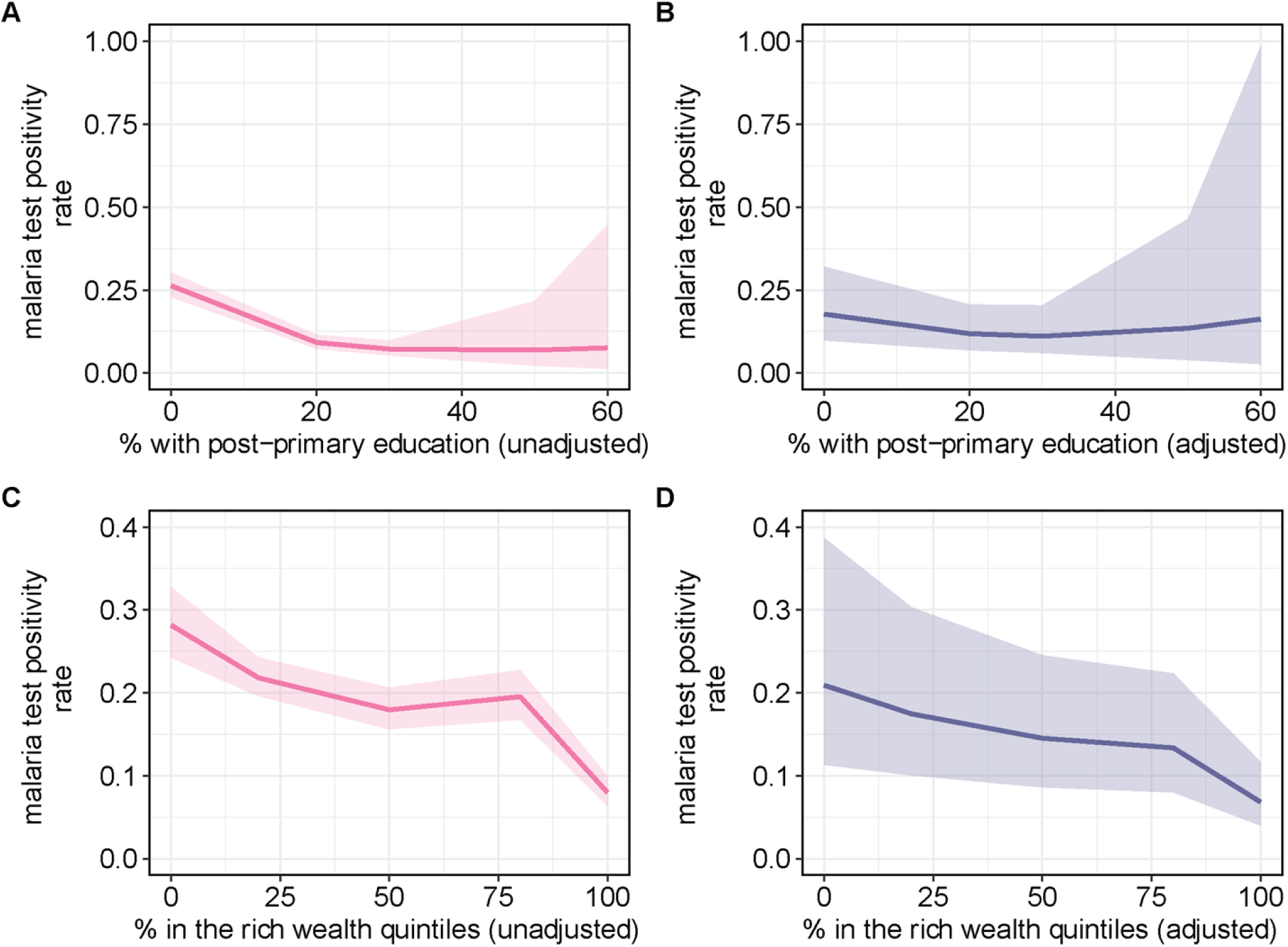
Effect plots of the bivariate and multivariate regression analysis for indicators of educational attainment and wealth. A) Unadjusted and B) adjusted effect of the percentage of individuals with post-primary education on malaria test positivity rate. Percentage of individuals with post-primary education was adjusted for percentage of individuals in the rich wealth quintiles, percentage of individuals living in improved housing in 2015, all age population density, median age, percentage of children under the age of five that sought medical treatment for fever, total precipitation and enhanced vegetation index. C) Unadjusted and D) adjusted effect of the percentage of individuals in the rich wealth quintiles on malaria test positivity rate. Percentage of individuals in the rich wealth quintiles was adjusted for percentage of individuals with post-primary education, percentage of individuals living in improved housing in 2015, all age population density, median age, percentage of children under the age of five that sought medical treatment for fever, total precipitation and enhanced vegetation index.

#### High population density and younger median age correlated with higher malaria transmission intensity

In the unadjusted analysis, malaria test positivity rate declined with increases in all age population density, up to 20,000 persons per square kilometer, before flattening out and going back up after 30,000 persons per square kilometer (Figure 5a). In the adjusted analysis, malaria test positivity rate remained mostly flat up to 20,000 persons per square kilometer after which increases in malaria test positivity was observed, albeit with high levels of uncertainity (Figure 5b). Declines in malaria test positivity rate followed increases in median age, particularly beyond median ages of 15 years old (Figure 5c - d).

**Figure 5:**
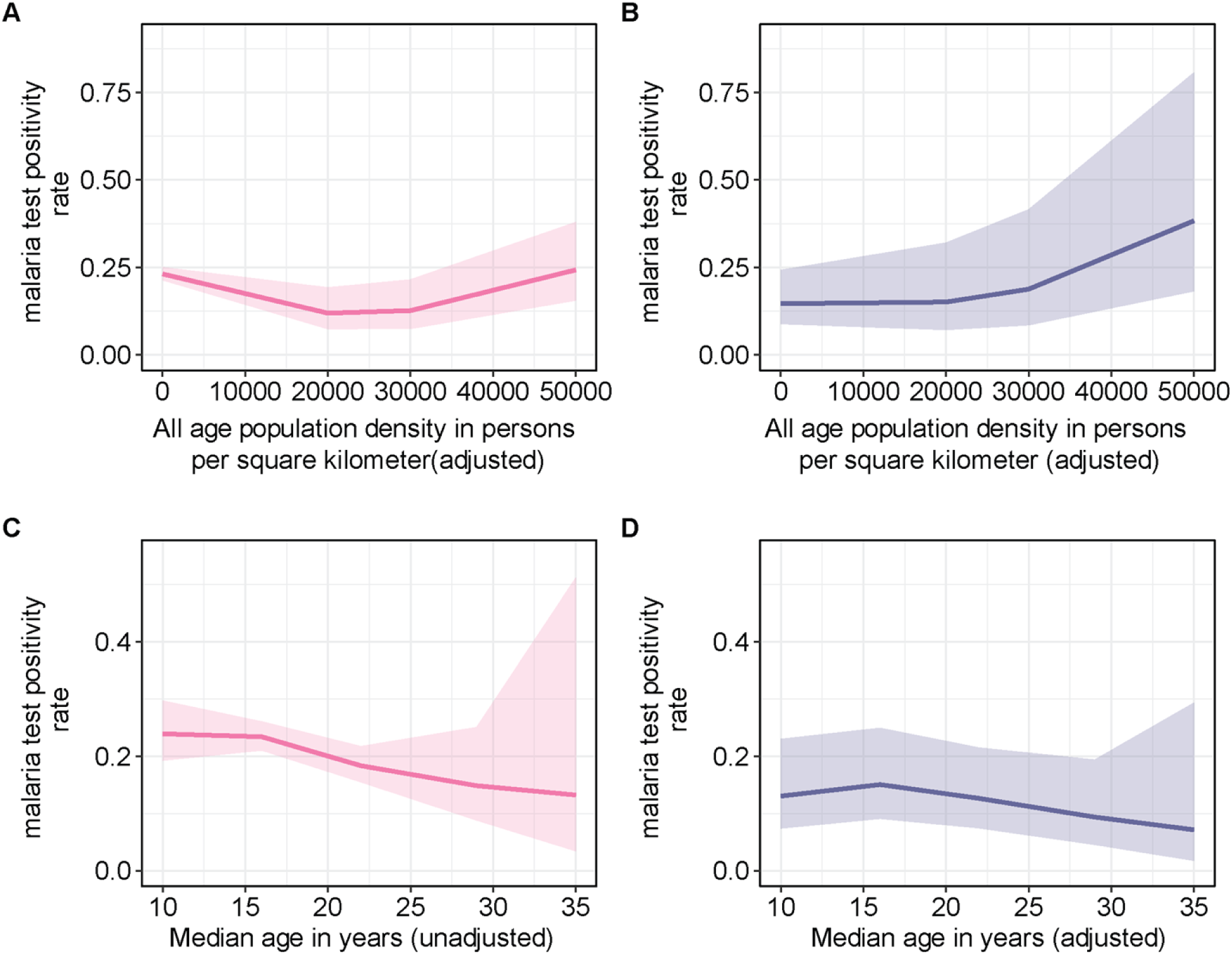
Effect plots of the bivariate and multivariate regression analysis for all age population density and median age. A) Unadjusted and B) adjusted effect of all age population density (persons per square kilometer) on malaria test positivity rate. All age population density was adjusted for the percentage of individuals with post-primary education, percentage of individuals in the rich wealth quintiles, percentage of individuals living in improved housing in 2015, median age, percentage of children under the age of five that sought medical treatment for fever, total precipitation and enhanced vegetation index. C) Unadjusted and D) adjusted effect of median age in years on the number of malaria test positivity rate. Median age was adjusted for percentage of individuals with post-primary education, percentage of individuals in the rich wealth quintiles, all age population density, percentage of individuals living in improved housing in 2015, percentage of children under the age of five that sought medical treatment for fever, total precipitation and enhanced vegetation index.

#### Higher enhanced vegetation index was positively associated with U5 malaria test positivity rate

In the unadjusted analysis, increasing malaria test positivity rate was correlated with increases in enhanced vegetation index, which characterizes vegetation cover and growth status. The greatest reductions in malaria test positivity rate, while highly uncertain, was observed at around vegetation indicies of 0.5 and 0.7 (Figure 6a). However, the effect of enhanced vegetation index was significantly diminished in the adjusted analysis (Figure 6b).

**Figure 6:**
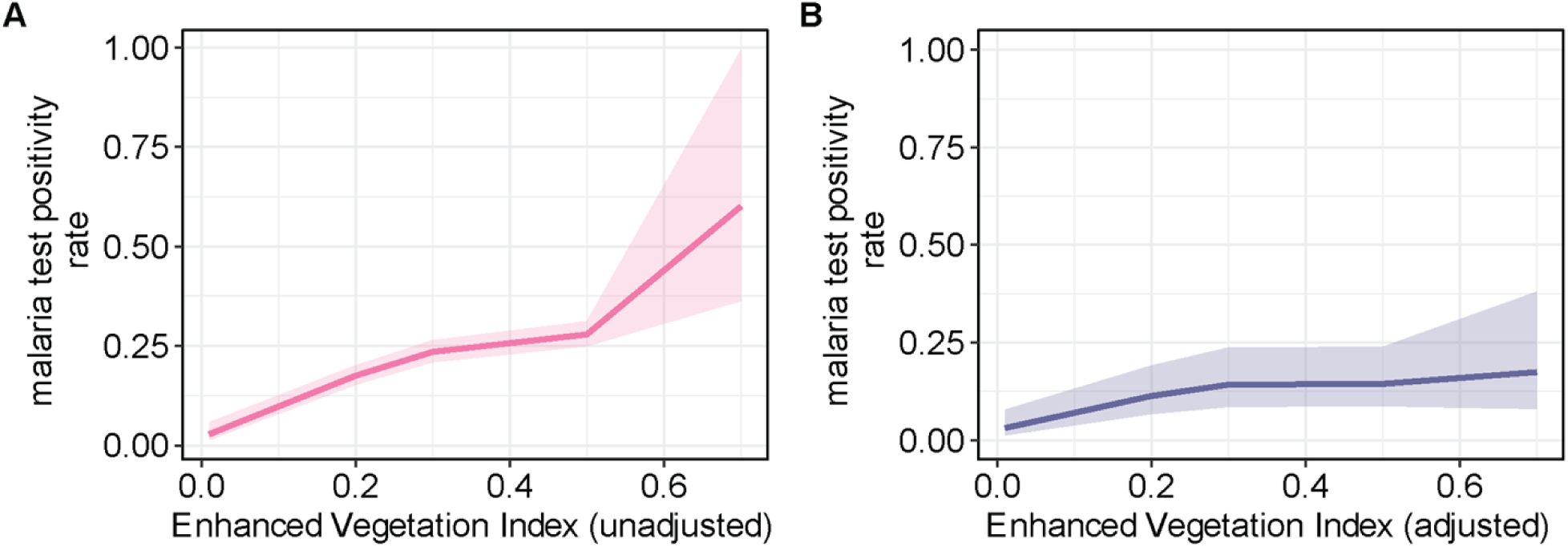
Effect plots of the bivariate and multivariate regression analysis for enhanced vegetation index. A) Unadjusted and B) adjusted effect of enhanced vegetation index on malaria test positivity rate. Enhanced vegetation index was adjusted for the percentage of individuals with post-primary education, percentage of individuals in the rich wealth quintiles, percentage of individuals living in improved in housing in 2015, all age population density, median age, percentage of U5 children that sought medical treatment for fever and total precipitation.

#### Effects of housing, care-seeking and precipitation

In the unadjusted analysis, increases in the proportion of individuals living in improved housing in year 2015 corresponded to very small increases in malaria test positivity rate, flattening out when improved housing proportions are between 40 to 60% after which a downward trajectory was observed (Figure 7a – b). Although the functional form of the association between the proportion of children under the age of five years that sought medical treatment for fever and malaria test positivity rate was nearly flat in both the adjusted and unadjusted analysis, the highest test positivity rate were observed at roughly between 20 and 50% (Figurer 7c – d). With regards to total precipitation during the month of the survey, the highest malaria test positivity rate were observed at 200 meters in the unadjusted analysis whereas the adjusted analysis, a clear negative relationship was observed (Figure 7e – f). Collectively, these findings suggests observed relationships may be due to the presence of information detection bias and/or residual confounding

**Figure 7:**
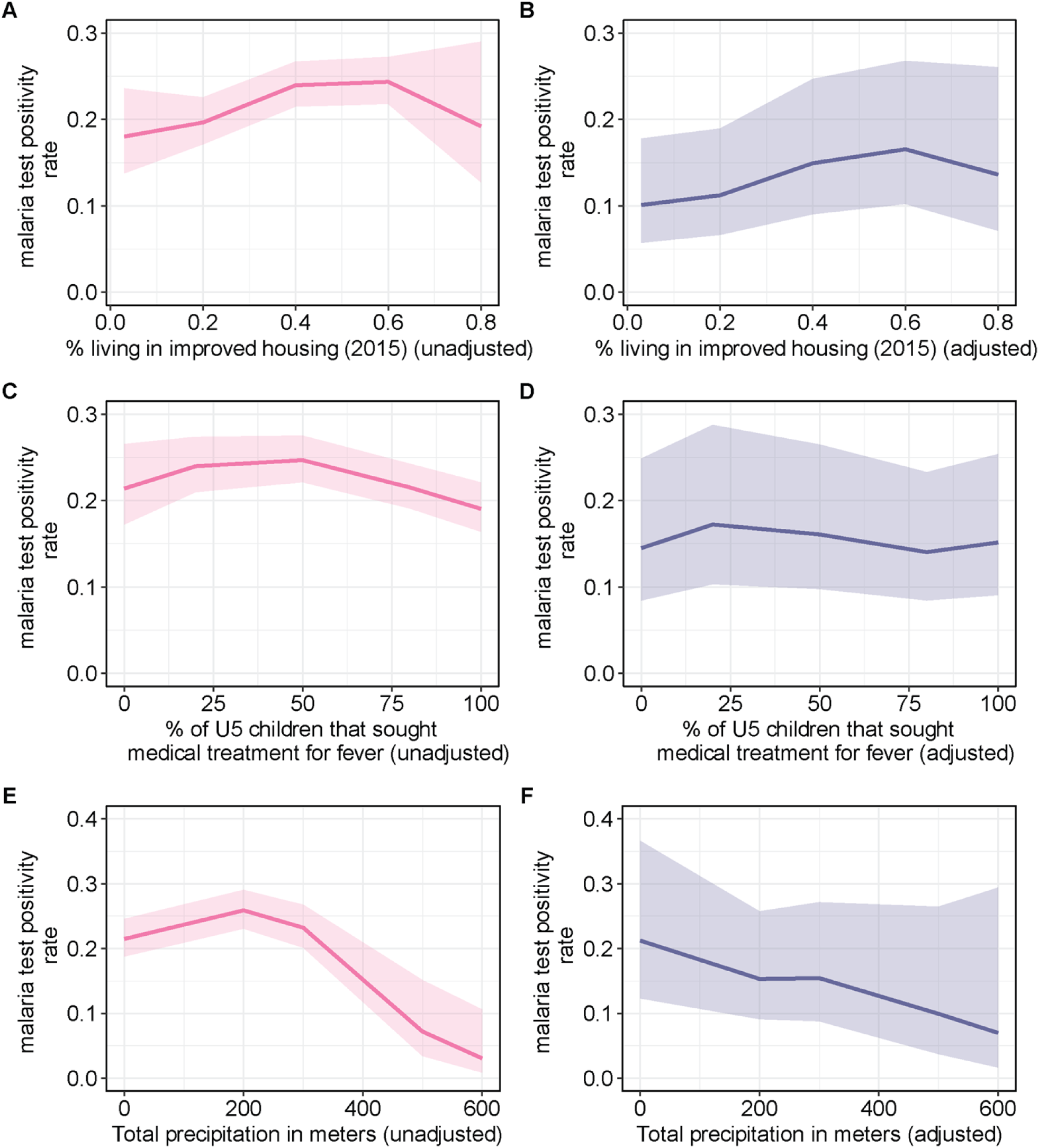
Effect plots of the bivariate and multivariate regression analysis for percentage of individuals living in improved housing in 2015, percentage of U5 children that sought medical treatment for fever and total precipitation. A) Unadjusted and B) adjusted effect of the percentage of individuals living in improved housing in 2015. Percentage of individuals living in improved housing in 2015 was adjusted for the percentage of individuals with post-primary education, percentage of individuals in the rich wealth quintiles, all age population density, median age, percentage of U5 children that sought medical treatment for fever, total precipitation and enhanced vegetation index. C) Unadjusted and D) adjusted effect of the percentage of U5 children that sought medical treatment for fever. Percentage of U5 children that sought medical treatment for fever was adjusted for the percentage of individuals with post-primary education, percentage of individuals in the rich wealth quintiles, all age population density, median age, percentage of individuals living in improved housing in 2015, total precipitation and enhanced vegetation index. E) Unadjusted and F) adjusted effect of total precipitation in meters. Total precipitation was adjusted for the percentage of individuals with post-primary education, percentage of individuals in the rich wealth quintiles, all age population density, median age, percentage of individuals living in improved housing in 2015, percentage of U5 children that sought medical treatment for fever and enhanced vegetation index.

## Discussion

We analyzed three years of the most recent DHS/MIS surveys to evaluate malaria burden among children the age of five years, identify predictors and generate effect plots with the potential to inform intervention prioritization using survey clusters administratively defined as urban areas.

U5 malaria test positivity rate in urban areas was found to be low, particularly in the North East and South South regions and to have declined over time. The low malaria test positivity rate observed in Nigeria’s North East may be due to the undersampling of the region as a result of insecurity. In addition, since the DHS/MIS survey outputs are typically unrepresentative of urban settings at the state and zonal level and the timing of the surveys fall within transmission months for states in the Southern geopolitical zones [3], we cannot make definitive conclusions of low malaria burden across urban settings or compare burden across geopolitical zones. To overcome temporal-related bias in comparisons in malaria test positivity across survey years, clusters sampled within the same months were compared but due to small sample sizes, we were unable to account for geopolitical zone-related differences in malaria seasonality.

The distribution of covariates obtained from the DHS/MIS are also likely unrepresentative of urban settings. For instance, educational attainment, as measured by the percentage of individuals with post-primary education (Secondary or college education) was low in majority of the sampled clusters whereas a greater proportion of clusters fell in the rich wealth quintiles or had improved housing infrastructure. Assuming that individuals correctly reported their educational attainment, it is improbable that most clusters will fall in the rich wealth quintiles or have improved housing infrastructure given the well-known positive correlation between educational attainment and these factors [46,47]. We attempt to overcome these limitations through the use of modeled covariates, where possible. However, we also provide visualizations of study covariates in the supplement to inform further study or data collection efforts.

The fitted lines depicting the relationship between U5 malaria test positivity rate and indicators of educational attainment, wealth, age distribution and vegetation cover and growth are aligned with findings from the existing literature [9,48]. In their summary of factors related to malaria transmission in urban areas across sub-Saharan Africa, Silvia and Marshall cited the literature showing that access to piped water, better refuse collection and improved access to prevention methods and treatment are among the numerous factors that reduce malaria risk among those of higher socioeconomic status in urban areas [9]. The higher risk of malaria infections among children has been previously described and be may be attributed to the higher malaria burden observed in clusters with more children [48]. Areas with high vegetation cover such as urban farms are well-known vector breeding sites [9]. However, the fitted line for the medical treatment seeking for fever were in discordance with the research literature [50], and suggest the presence of residual confounding or information detection bias. Additional investigations is essential to understand the sources of confounding and bias related to treatment seeking.

Despite the aforementioned data gaps, the methods from this study can be applied to local data to inform stratification interventions in urban areas. The multivariable model can be used to predict areas with high U5 malaria burden and explore the causal impact of various factors. The methods used for generating the functional forms of the bivariate association with predictors can be helpful in selecting thresholds for intervention prioritization and deprioritization. For instance, an effect plot that shows that urban localities where < 20% of residents have post-primary education are at high malaria risk could be used to inform prioritization of vector control tools to these areas, if transmission is believed to be local or for the provision of prophylaxis if transmission is heavily influenced by mobility patterns. Improved data that adequately captures malaria burden and related sociodemographic and environmental factors is required for the application of the study methologies to meaningfully inform intervention prioritization and deprioritization decisions.

The bivariate analysis provides several insights that require further exploration. Among them is the observed regional differences in bednet use. We found that U5 malaria test positivity rate was highest among clusters with the lowest bednet use in the Northern geopolitical regions whereas in the Southern regions, malaria test positivity rate appeared to be uniform in the South West region irrespective of bednet use or higher among those that use nets as in the South-East region (Supplementary figure 18a). Further study is needed to clarify why the effects of net use would vary by geopolitical region. Additionally, further research is needed to understand the role of employment type on community malaria transmission. Using the 2018 DHS, which was the only survey year with employment type, we found that increases in the cluster proportions of male partners who are agricultural workers correlated with small increases in the number of malaria positives in children (Supplementary figure 12a). Routine surveillance data has higher temporal resolution compared to survey data and associated data collection systems can be used to collect data on how the distribution of different parental employment types impact malaria transmission among children residing in urban settings. Understanding regional variance in bednet use and the role that employment is important for the design of contextually relevant malaria interventions.

This study complements the existing research literature by describing spatial-temporal variations in U5 malaria test positivity and associated risk factor relationships in urban areas within Nigeria. Efficiency in intervention distribution could potentially lead to reductions in malaria burden and when locally representative data is available for individual cities, study methods can be applied to inform intervention stratification strategies. With increasing urbanization, there is growing interest to address malaria in urban areas as evidenced by the recent convening of experts by the WHO as part of a technical consultation on the burden and response to malaria in urban settings. By highlighting the data gaps inherent in the Nigerian DHS/MIS, we hope that this encourages additional investments to improve its design and its utility, as well as in routine surveillance systems.

## Data Availability

Raw DHS/MIS data can be downloaded after approval by the DHS program from https://dhsprogram.com/. Data summaries and code used to construct statistical model in this study are publicly available via the following doi: 10.5281/zenodo.6350331

10.5281/zenodo.6350331

## Acknowledgements

**None**

## Supporting Information

**S1 File: Supplementary appendix**

